# Parenthood, Caregiving and Female Fertility Among General Internal Medicine Physicians in Switzerland

**DOI:** 10.1101/2025.09.10.25335526

**Authors:** Isaac Egger, Susanna Weidlinger, Jarmila Zdanowicz, Karolina Kublickiene, Christa Nater, Sven Streit, Jeanne Moor

## Abstract

**Background/Importance:** Balancing work and family life is a particular challenge for physicians, yet finding a good balance is crucial given that childcare responsibilities and institutional support strongly influence career trajectories. This study examined female fertility and gender differences in family founding, working hours and childcare responsibilities among physicians in Switzerland.

**Objective:** Characterize fertility and family-founding among women physicians, compare women’s and men’s family-founding attitudes, and examine how work patterns, caregiving responsibilities and perceived institutional support relate to reproductive outcomes.

**Design, Setting, and Participants:** Secondary analysis of an existing, anonymous, cross-sectional, web-based survey of the Swiss internal medicine workforce (fielded from December 2021 to April 2022).

**Main Outcomes and Measures:** Desire to have children, past or current intent to delay childbearing, self-reported female infertility, number of children, maternal age at first birth, perceived adequacy of external childcare, and employer respect of pregnancy-related working-hour limits. Group comparisons used Welch’s t-tests and chi-square tests, and associations with workload used linear regression models.

**Results:** This analysis included 682 physicians, 278 (41%) men and 404 (59%) women. Men’s age was 39±12 years (mean ± standard deviation [SD]), and women’s age was 36±10 years. The prevalence of self-reported infertility among female physicians was 27.7%, which exceeds Swiss general population estimates of 10-15% (p<0.001). The mean age at first birth among physician-mothers was 31.3 years, which did not differ from the Swiss average (p=0.388). Women physicians had fewer children than men physicians (p=0.006). Among physicians without children, more women (69.8%) than men (51.6%) currently postponed having children (p<0.001). Also, among parents more women (42.2%) than men (21.8%) had delayed having children (p<0.001). Overall, satisfaction with childcare was limited, with 37% of men and 30% of women rating their childcare situation as inadequate. Among women who had ever been pregnant during employment (n = 123), 57% reported that legally prescribed pregnancy-related working-hour limits were not respected.

**Conclusions and Relevance:** In this national physician sample, women reported having fewer children and more frequently postponing having children than men, and experienced a higher prevalence of self-reported infertility than the Swiss population. Perceived childcare adequacy was suboptimal for both genders, and many reported insufficient adherences to pregnancy-related working-hour protections. Improving scheduling, enforcing maternity protections, increasing access to reliable childcare, and promoting equitable caregiving norms could enhance reproductive and professional outcomes for women in medicine in Switzerland.

## Introduction

Although women now comprise a growing share of the medical workforce, they frequently delay childbearing and face unique barriers during training and in the early stages of their careers. Large studies show that physicians tend to start families later than nonphysicians (median age at first birth: approximately 32 vs. 27 years), with a skewed distribution toward older maternal ages.^1^ In Switzerland, approximately 90% of women intend to have at least one child, and the mean maternal age at first birth is 32 years; nevertheless, nearly one quarter of women remain.^2^ Infertility affects an estimated 10 - 15% of couples in Switzerland, which is broadly consistent with global lifetime prevalence estimates ranging from 10.7% in the Eastern Mediterranean to 23.2% in the Western Pacific region.^3^

Evidence from the United States and Europe indicates that women physicians face a particularly high burden, with infertility rates of 25–37%—nearly double those of the general population - and a markedly higher likelihood of undergoing fertility treatment.^4,5^ Surgical and other procedure-intensive specialties report elevated infertility treatment use and pregnancy loss compared to the general population.^5^ Contributing factors include long and irregular hours, missed prenatal care, night-shift work, physical and psychological demands, exposure to potentially embryo-/fetotoxic situations and substances (ionizing radiation, anesthetic gases, surgical smoke, etc.) as well as cultural and structural barriers, such as limited parental leave and inadequate lactation and childcare support.^6,7^

Despite growing awareness of these issues, there is limited European data, and Swiss physicians are underrepresented in existing research.

General Internal Medicine (GIM) physicians represent the largest medical specialty in Switzerland, with a high proportion of women and a central role in primary and hospital care, making GIM particularly relevant for studying gender-specific challenges in family planning and career development.

The present study aimed to address this knowledge gap by analyzing nationwide survey data from Swiss physicians specializing in internal medicine. Specifically, our objectives were to: (1) describe fertility and family-founding patterns among female physicians, (2) compare selected outcomes with those of male physicians, and (3) examine work patterns, caregiving, and perceived support relevant to reproductive outcomes. We place our findings in context with contemporary literature on physician fertility, pregnancy risks, and workplace exposures.

## Methods

### Design

This secondary analysis is based on an anonymous, web-based cross-sectional survey that was originally designed to evaluate the career ambitions and well-being of Swiss internal medicine physicians.^8,9^

### Participants

Physicians in General Internal Medicine in Switzerland were invited to participate through 14 Swiss hospitals, six ambulatory/primary care institutes, and two newsletters circulated by the Swiss Society of General Internal Medicine (SGAIM) and the Swiss Young Practitioners’ Association (JHaS). Additionally, a free advertisement in the Swiss medical journal Primary and Hospital Care invited participation.

### Survey

The survey was hosted on SurveyMonkey from December 2021 to April 2022. Respondents provided electronic informed consent for voluntary participation. The local ethics committee of the Canton of Bern waived formal review (ID: Req-2021-0108).

### Variables

The survey^8^ collected demographic data, as well as data on personal priorities, well-being, family and reproductive status, professional environment, career ambitions, and work activities. For the present analysis, we focused on the following:

1. Reproductive and family-founding variables, including desire to have children, past and current intent to delay having children, parity, and age at first birth among physician-mothers.
2. Female infertility (self-reported): defined as failure to achieve a pregnancy after 12 months of regular unprotected intercourse in women <35 years, or after 6 months in women ≥35 years when trying to conceive.^10,11^
3. Work and caregiving context: specialty grouping (e.g., general internal medicine vs. non-GIM), work fraction (“work quota,” percentage of full-time equivalent [FTE]), weekly work hours, on-call/irregular hours, where available, perceived adequacy of external childcare (Likert-type scale), and employer respect of pregnancy-related working-hour limits (Likert-type scale).

To provide context, we obtained estimated reference values from the Swiss general population (maternal age at first birth and infertility prevalence) from national data and published reports.^12–14^

### Study outcomes

The primary outcomes were the gender-specific frequencies of the following: (i) wanting to have children, (ii) past or current intent to delay having children, (iii) female infertility as defined above, and (iv) maternal age at first birth.

Secondary analyses examined work quotas, weekly work hours, perceived adequacy of external childcare, and, where applicable, employers’ respect for pregnancy-specific working-hour limits.

### Statistical analysis

An a priori power calculation was not performed for this secondary analysis. Descriptive statistics were used to summarize infertility prevalence, family-planning intentions, and age at first birth. Continuous variables are presented as the mean and standard deviation, or as appropriate, as distributions. Categorical variables are presented as counts and percentages. We used Welch’s t-tests for continuous variables that were approximately normally distributed and chi-square tests or Fisher’s exact tests as needed for categorical variables to make between-group comparisons (women vs. men). Informative sex-stratified age structures were visualized by population pyramids where applicable.

To explore the associations between gender and workload measures (e.g., weekly hours, employment fraction), we fitted univariable linear regression models. All tests were two-sided with α = 0.05, and no adjustments for multiplicity were applied. Analyses were conducted in R version 4.3.2 (R Foundation for Statistical Computing, Vienna, Austria).

## Results

This study included 682 physicians. Among them, 278 (41%) were men and 404 (59%) were women. The mean age of the men was 39±12 years, and the mean age of the women was 36±10 years (mean ± standard deviation [SD]) (Table 1).

**Table 1.** Population characteristics

| **Table 1 Population characteristics** |  |  |  |  |
| --- | --- | --- | --- | --- |
| **Physicians** | **Male physicians** |  | **Female physicians** |  |
|  | **N=278** |  | **N=404** |  |
| **Age in years, mean (SD)** | 39 (12) |  | 36 (10) |  |
| **Swiss language region** |  |  |  |  |
| **German-speaking** | 197 | 71% | 288 | 71% |
| **French-speaking** | 73 | 26% | 111 | 27% |
| **Italian-speaking** | 8 | 2.90% | 5 | 1.20% |
| **Type of workplace** |  |  |  |  |
| **University hospital** | 104 | 37% | 155 | 38% |
| **Other hospital** | 115 | 41% | 152 | 38% |
| **Private practice with ≥3 colleagues** | 23 | 8.30% | 43 | 11% |
| **Private practice with <3 colleagues** | 29 | 10% | 36 | 8.90% |

### Infertility

The self-reported infertility prevalence among female physicians was 27.7%, which is higher than the estimated 10-15% prevalence of female infertility in the general population of Switzerland (Table 2) (single-sample chi-squared test, p<0.001).^13,14^

**Table 2:** Comparison of study population with Swiss general population

| Table 2: Comparison of study population with Swiss general population |  |  |  |
| --- | --- | --- | --- |
| Outcome | Swiss general population in 2022 | Female physicians | p-value |
| Infertility prevalence | 10-15% | 28% | <0.001<sup>1</sup> |
| Age at birth of first child | 31.3 years | 31.0 +- 3.9 years* | 0.388<sup>2</sup> |
| <sup>1</sup>One-sample Chi-square test, assuming a population prevalence of 12.5%. <sup>2</sup>One-sample t-test.<br>*mean +- standard deviation. |  |  |  |

### Desire to have children

Overall, the prevalence of already having or desiring to have children or already having children was high among physicians (Figure 1B). At the beginning of their careers, under the age of 30, 22% of male and 33% female physicians were uncertain whether they wanted to have children in the future (p=0.004). This uncertainty decreased with age (Figure 1C).

**Figure 1.**
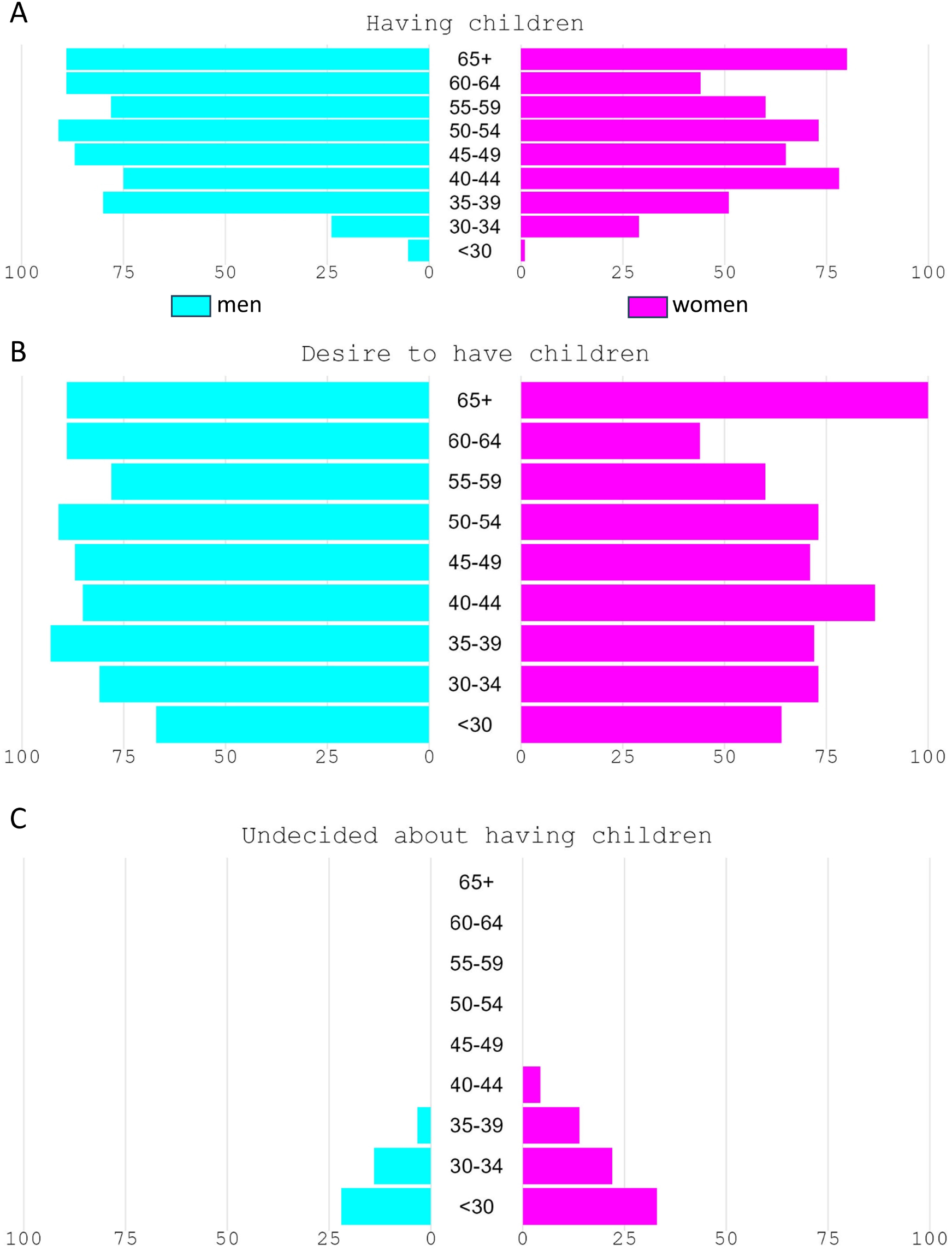
The Percent of physicians who have or want to have children, and the number of undecided individuals. X-axis depicts percent.

### Parenthood and age at first birth

Figure 1A shows that female physicians were more likely to be childless than their male colleagues (67% vs. 52%, p<0.001). Female physicians also had fewer children than male physicians (p<0.001) (Supplemental Figure 1). The age distribution at first birth among physician-mothers is presented in Supplemental Figure 2.

### Intent to Delay Childbearing

Gender differences were observed in both past and current intentions to delay having children. Figure 2A shows that depending on age group, between 10% and 25% of male physicians reported to have postponed starting a family. However, the proportion of female physicians who stated that they had postponed starting a family was twice as high in most age groups. Among physicians without children, 69.8% of female physicians and 51.6% of male physicians are postponing having children (p <0.001 for gender difference). Among physicians who already have children, 42.2% of female physicians and 21.8% of male physicians stated that they had postponed starting a family (p <0.001 for gender difference), and about two thirds of each gender considered their childcare situation as adequate (Supplemental Table 1).

**Figure 2.**
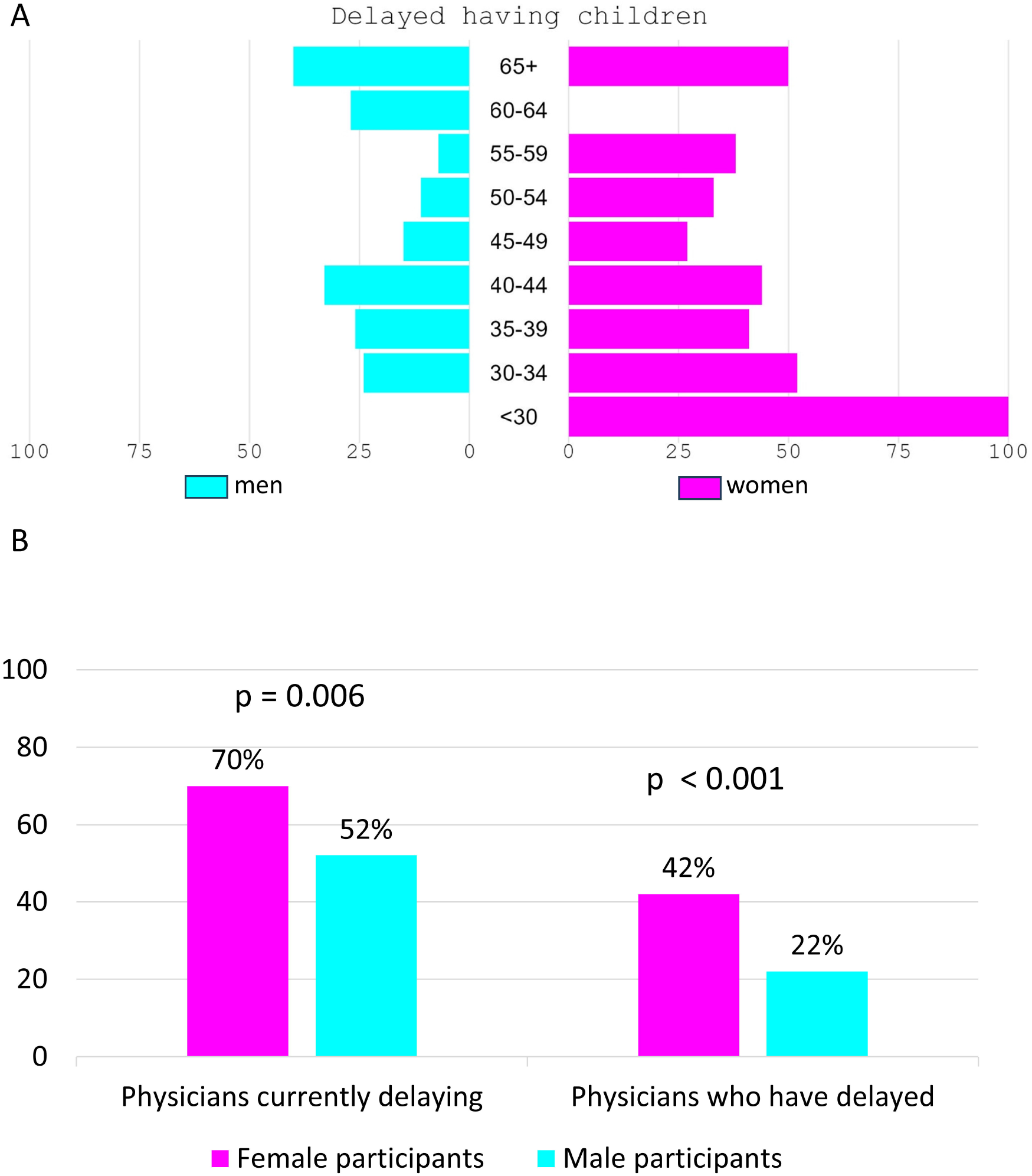
The fraction of physicians (A-B) or medical students (B) who have delayed or are currently delaying having children. The x-axis in panel A depicts percent.

### Gender differences in domestic responsibilities

Figure 3A shows that, among physicians who became parents, women worked significantly fewer hours per week than men after the birth of a child. This gender difference in weekly working hours persisted for several years until the children were aged 13–18. Similar data was obtained for the work quota: While the work rate of non-parents remained at 100%, the work rate of female physicians with children was significantly reduced after childbirth, whereas the work rate of male physicians remained unaffected (Figure 3B). This gender-specific difference in work rate only disappeared once the children were over 18 (Figure 3B).

**Figure 3.**
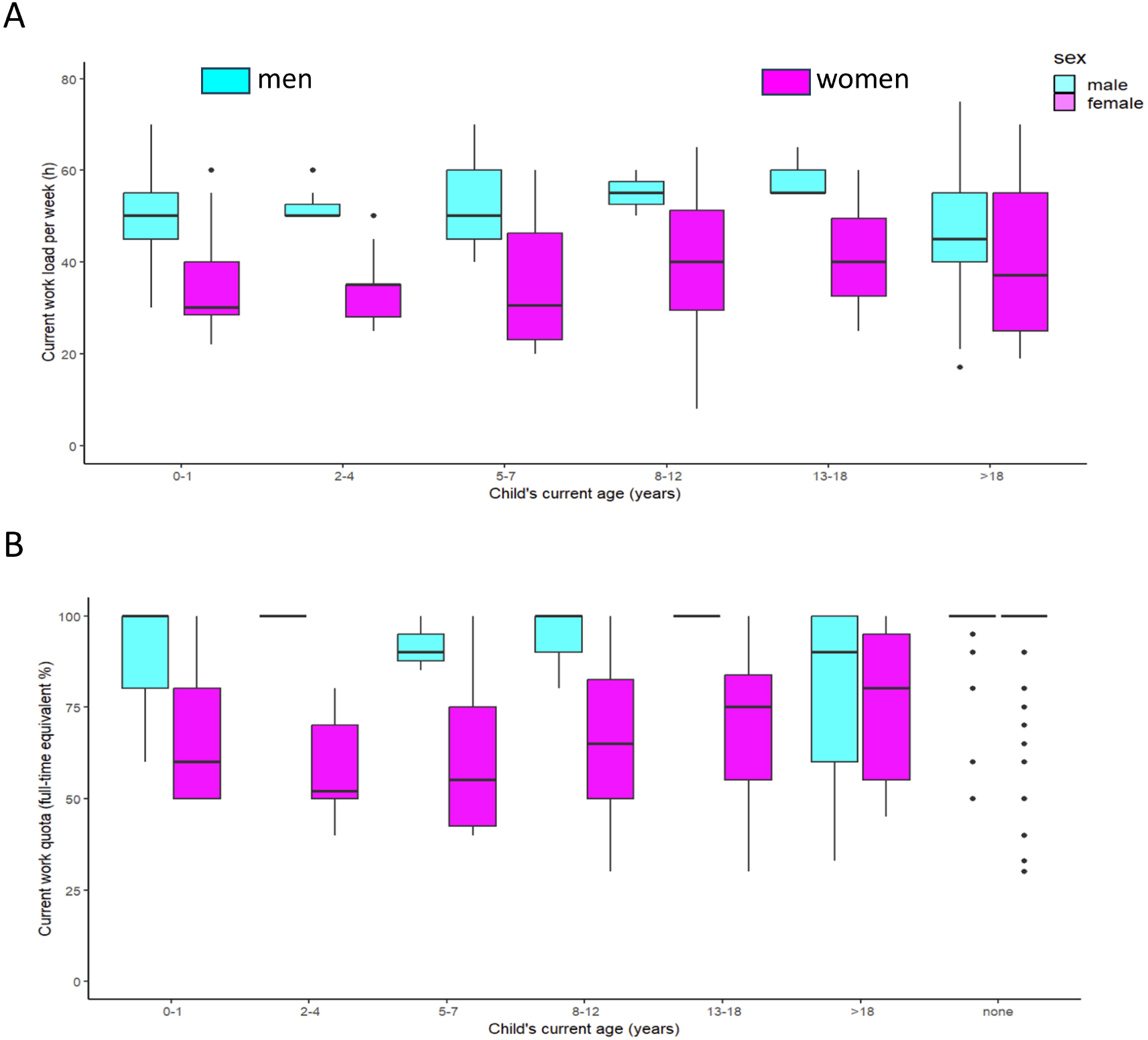
Female physicians are more likely than male physicians to act as the primary caregiver for their young children than male physicians, according to working hours (A) and work quota (B). P < 0.001 for the sex effect in both A and B.

Self-reported participation in childcare tasks among assistant physicians showed a comparable distribution between the sexes (Figure 4B). However, from the level of senior physicians onwards, the self-reported share of childcare and household tasks was substantially greater for female physicians than for their spouses (Figures 4B–C). For example, the n=18 male and n=7 female senior physicians in GIM with available data reported median shares of 42% (IQR: 42-50%) and 60% (IQR: 50-75%) for childcare tasks respectively, while n=27 male and n=44 female GIM attending physicians reported 40% (IQR: 30-50%) and 60 % (IQR: 50-70%), whereas n=71 male and n=122 female GIM residents reported 50% (IQR: 40-50%) and 50% (IQR: 40-65%), respectively. (Figure 4B). Similar data were provided for household task share (Figure 4C).

**Figure 4.**
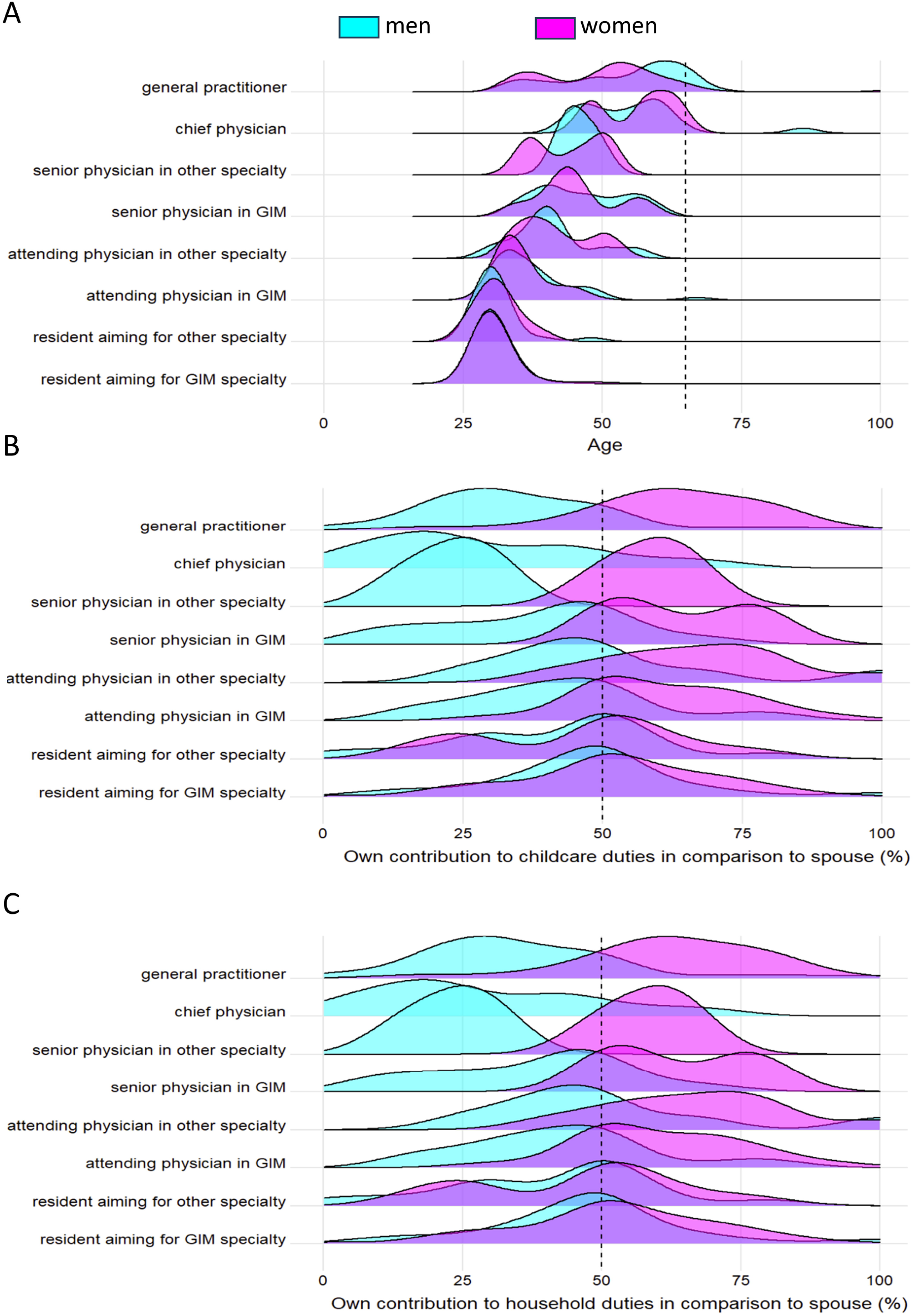
After residency, female physicians contribute more to childcare and household duties than male physicians. Figure A shows the age distribution, and Figures B and C show the self-reported contribution to childcare and household duties for all participating physicians, stratified by sex, at different hierarchical positions. Vertical lines indicate retirement age (A) and 50% of domestic contribution (B-C).

### Employer Respect of Pregnancy-Related Working-Hour Limits

Finally, compliance with legally prescribed working time limits during pregnancy was reported to be insufficient by employers. Among respondents who had been pregnant during employment (n=123), 57% reported working-hour limits were “never/mostly not/rather not” respected, versus 44% reporting “rather/mostly/always” respected. This highlights a significant discrepancy between legislation and practice. (Supplemental Table 2).

## Discussion

In a large Swiss sample of physicians, female physicians reported having fewer children, shouldering a heavier domestic care burden, and often experiencing the inadequate implementation of pregnancy-specific working time protection measures compared to male physicians. These patterns align with U.S. findings on delayed family formation, increased use of fertility treatments, and workplace barriers during medical training and early careers.^1,5^ In the following sections, we discuss the key findings with their implications for the healthcare workforce.

### 1. Fertility

One of the most striking findings of this study is the high prevalence of self-reported infertility among female physicians (27.7%), nearly double the prevalence estimated in the Swiss general population. This observation mirrors US and European reports showing infertility rates of 24 - 43% among women physicians.^4,5,15^ Several explanations are plausible, including delayed childbearing, occupational exposures (radiation, anesthetic gases, night shifts), and lifestyle factors such as insufficient sleep and chronic stress. Physicians also face higher rates of pregnancy complications, including miscarriage, gestational diabetes, and hypertensive disorders of pregnancy.5,16 These findings suggest that fertility itself should be recognized as a central dimension of gender inequity in medicine, and targeted interventions - such as fertility education during medical training - may help mitigate risks.

### 2. Delayed Parenthood and Reduced Family Size in Female Physicians

Female physicians not only report intentions to postpone parenthood but also actually enter into parenthood at significantly older ages and have fewer children than their male colleagues. This aligns with findings from the SwissMedCareer Study, which showed that female physicians—especially those with children - experience slower career advancement and often opt for part-time work or career breaks to accommodate family life.^17,18^ Such patterns reflect broader structural mechanisms contributing to the “child penalty” phenomenon marked by long-term setbacks in women’s labor market outcomes after childbirth.^19^ Beyond structural and cultural barriers, occupational lifestyle factors may also play an important role. Long working hours, irregular schedules, and night shifts are associated with disruption of circadian rhythms, which has been linked to subfertility and adverse obstetric outcomes.^20^ In Switzerland, one-third of primary care providers report insufficient sleep duration,^21^ and excessive workloads are common among hospital-based physicians.^22^ Chronic stress, with its impact on the hypothalamic-pituitary-gonadal axis, represents an additional modifiable pathway by which physician working conditions could impair fertility and pregnancy outcomes.^23^

### 3. Poor Compliance with Pregnancy Work Protections

Our findings raise concern: only a minority of employers fully adhered to legally mandated working-hour protections for pregnant physicians. This echoes reports of heterogeneous implementation of maternity protection legislation (OProMa) in Swiss workplaces, particularly within healthcare settings.^24–26^ The lack of systematic application of protective measures such as risk analyses or adapted job roles undermines both maternal well-being and gender equity.

### 4. Inadequate Childcare and Unequal Household Burden

A third of both female and male physicians considered their childcare situation as inadequate, with female physicians contributing a larger share of domestic work and often taking on the role of primary caregiver, particularly so post-residency. These findings echo prior literature and represent the so-called “second shift” effect, wherein professional women continue to bear most of domestic labor.^27,28^ In Switzerland specifically, the OECD has highlighted structural barriers such as irregular school hours, high childcare costs and insufficient family-friendly policies that exacerbate work-family conflicts especially for mothers, hinder gender equity, and diminish the market participation of skilled mother-workers.^29,30^

### 5. Cumulative Career and Well-being Consequences

Taken together, delayed parenthood, fewer children, weak enforcement of pregnancy protections, and caregiving burdens form a cumulative disadvantage that likely hinders career trajectories, increases burnout risk, and may lead to attrition among female physicians. These dynamics echo older Swiss findings where physician mothers advanced more slowly, had less career support, and often reduced their employment status.^17,18^

### 6. Implications for Policy and Practice

To address these inequities, a comprehensive strategy is essential:

i. **Strengthen enforcement of maternity protections:** Ensure universal compliance with OProMa (*Ordonnance sur la protection de la maternité au travail*) by incorporating workplace risk analyses and timely protective measures.
ii. **Expand affordable, accessible, and reliable childcare:** Facilitate better professional continuity, especially for families juggling clinical duties.
iii. **Promote cultural change:** Normalize sharing of caregiving and domestic responsibilities across genders through institutional role modeling and supportive policies.
iv. **Implement targeted career support for women:** Mentoring programs, flexible career structures, and part-time or phased career pathways can counteract the “child penalty” and retain female talent in medicine.
v. **Integrate fertility education into medical training:** According to prior research, providing education on fertility and the risks of delayed childbearing during medical school or residency could mitigate some of these challenges.^31^ Combined with institutional flexibility, such as job-sharing, clear parental leave policies, and part-time options, these initiatives could better support physicians who are planning families.

### Limitations

This study has several limitations. First, infertility was assessed only through self-reporting among women, and male physicians were not asked about infertility or pregnancy complications experienced by their partners. Second, detailed data on biological and lifestyle risk factors (e.g., smoking, body mass index [BMI], and stress) were unavailable because the original survey was not designed for this purpose. Third, voluntary participation may have introduced response bias, as physicians with a greater personal interest in family building or reproductive health may have been more likely to participate. Fourth, the cross-sectional design precludes causal inference, so the observed associations between gender, workload, and fertility outcomes should be interpreted descriptively. Finally, comparisons with the general Swiss population relied on published statistics rather than contemporaneous non-physician control groups.

### Conclusion

The present data provide first evidence of a widespread lack of accommodation for family-founding among General Internal Medicine physicians in Switzerland, which is associated with delaying having children especially among female physicians, lower numbers of children among female compared to male physicians, or even female infertility. Identifying and overcoming the potential underlying reasons such as traditional gender roles, rigid work quota expectations and inadequate childcare options and may be pivotal to sustain reproductive health of the internist healthcare workforce.

## Acknowledgements

The authors thank the respondents, and the Swiss Society of Internal Medicine, the Swiss Young General Practitioners Association (JHaS), and the hospitals involved with distribution of the survey.

## Funding

The analyzed survey study was funded by the Swiss Society of Internal Medicine. No additional funding was obtained for the present analysis.

## Conflicts of interest

All authors have no conflicts of interest to declare.

## Data availability

The raw data from the analyses presented in this study are available upon reasonable request to the authors.

## Notes

### Competing Interest Statement

The authors have declared no competing interest.

### Author Declarations

The ethics committee of the Canton of Bern waived this study (ID: Req-2021-0108).

